# Distribution of Incubation Period of COVID-19 in the Canadian Context: Modeling and Computational Study

**DOI:** 10.1101/2020.11.20.20235648

**Authors:** Subhendu Paul, Emmanuel Lorin

## Abstract

We propose an original model based on a set of coupled delay differential equations with fourteen delays in order to accurately estimate the incubation period of COVID-19, employing publicly available data of confirmed corona cases. In this goal, we separate the total cases into fourteen groups for the corresponding fourteen incubation periods. The estimated mean incubation period we obtain is 6.74 days (95% Confidence Interval(CI): 6.35 to 7.13), and the 90^th^ percentile is 11.64 days (95% CI: 11.22 to 12.17), corresponding to a good agreement with statistical supported studies. This model provides an almost zero-cost approach to estimate the incubation period.

## Introduction

The outbreak of coronavirus disease 2019 (COVID-19), first appeared in Wuhan (China) and spread around the world (*20*), and is creating dramatic and daily changes with profound impacts worldwide. People with underlying medical condition, respiratory disease, diabetes, cancer etc., and older people are vulnerable to severe complications and death from coronavirus, although we are discovering new features of COVID-19 every day. In the absence of vaccination and proper medication, we can merely obey some non-pharmaceutical containments, lockdown, social distancing, hand hygiene, face masking, mobile app to trace corona-positive individuals, to help prevent the spread of the this infectious disease. To minimize transmission of the virus through human-to-human interaction, quarantine of individuals with exposure to infectious pathogen is an effective strategy for containing contagious diseases. To govern a quarantine period for asymptomatic as well as presymptomatic individuals, it is essential to fully estimate the incubation period of COVID-19 disease.

*The incubation period* of an infectious disease is the *time between when a person is infected by a virus and when the first symptoms of the disease are noticed*. Estimates of the incubation period for COVID-19 range from 2 to 14 days, according to the investigation so far. Precise knowledge of the incubation period is crucial to control infectious disease like COVID-19; a long incubation period means a high risk of further spreading the disease. The distribution of the incubation period can be used to estimate the basic reproduction number *R*_0_, a key factor of epidemics, in order to measure the potential for disease transmission. It is indeed difficult to obtain a good estimate of the incubation period on the basis of limited data.

There are several statistical studies (*3, 4, 9, 11–15, 17, 23, 28*), using a single measure (*17*), estimating the incubation period of the current pandemic. In addition to those statistical approaches, there are numerous analytical and computational studies based on mathematical models, involving Ordinary Differential Equations (ODE) (*5, 8, 10, 19, 21*) as well as Delay Differential Equations (DDE) (*6, 7, 18, 22, 26, 27*), to calculate the basic reproduction number *R*_0_ and understand the underlying dynamics of the epidemic. Researchers usually consider a single delay models, occasionally two delays.

To the best of our knowledge, we are proposing for the first time a mathematical model, comprising fourteen delays, to estimate the incubation period utilizing publicly available data of the total number of corona-positive cases. This approach is free from any special type of samples in order to produce the distribution of the incubation period. It is then almost cost free, as it only involves a small scale computations. After a single calculation employing this method, we can generate the current distribution as well as previous distributions of the incubation period. We can also observe the change in the incubation period. In the statistical based approach, it is usually difficult to consider a large incubation period if the sample size is small. However, in this approach, we can go well beyond 14 days, the incubation period we have set for the current work. In this context, we demonstrate the incubation period of the COVID-19 epidemic in Canada employing publicly available data of confirmed corona-positive cases (*1*). As of November 7, 2020, the World Health Organization (WHO) had confirmed a total of 251,338 cases of COVID-19 in Canada, including 10,381 deaths (*20*).

There are several studies on incubation period mainly based on Chinese patients that can only provide a rough estimation for rest of the world. The incubation period may depends on age (*25*) (median-age */* country), hard immunity, public health system, corona testing capacities, daily corona cases, etc. For a better estimation of the incubation period for a particular region, we need to study local patients. Data collection is a bottleneck in studying the incubation period. However, one can easily estimate the incubation period using the approach we propose and publicly available data of confirmed cases.

## METHODS

In this section, we introduce a compartment based infectious disease model including a total of seventeen partitions, Lockdown, Susceptible, Infected and fourteen compartments of Total confirmed cases (LSIT). The model is constructed as a set of coupled delay differential equations involving several variables and parameters.

### The Model

Modeling the spread of epidemics is an essential tool for projecting its outcome. By estimating important epidemiological parameters using the available database, we can make forecasts of different intervention scenarios. In the context of compartment based model, where the population of a region is distributed into several population groups, such as susceptible, infected, total cases etc., is a simple but useful tool to demonstrate the panorama of an epidemics.

In this article, we introduce a infectious disease model, extending the standard SIR model, including the phenomenon lockdown, a non-pharmaceutical way to prevent the spread of the epidemics. The schematic diagram of the model is presented in **Fig. S1** with several compartments and various model parameters. The following are the underlying principles of the present model.

- The total population is constant (neglecting the migrations, births and unrelated deaths) and initially every individual is assumed susceptible to contract the disease.
- The disease is spread through the direct (face-to-face meeting) or indirect (through air current, common used or delivery items like door handles, grocery products) contact of susceptible individuals with the infective individuals.
- The quarantined area or the compartment for corona cases contains only members of the infected population who are tested corona-positive.
- The virus always kills some percent of the people it infects; the survivors percent represents the recovered group.
- There is a non-pharmaceutical policy (stay at home), commonly known as *lockdown*, to stop the spread of the disease.

Based on the above principles, we consider several compartments:

- Susceptible (*S*): the group of individuals who can be infected.
- Infected (*I*): the group of people who are spreading the contiguous disease.
- Total cases (*T*): the group of individuals who tested corona-positive (Active cases + Recovered + Deaths).
- Lockdown (insusceptible) (*L*): the group of persons who are keeping themselves safe.

The goal of the present model is to estimate the distribution of the incubation period of COVID-19. In this goal, we split the compartment *T* into *J* subcomponents *T*_1_, ⋯, *T*_J_, where

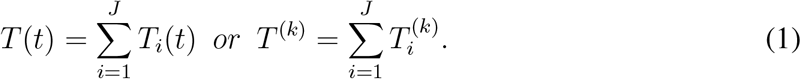

In **Eqn. 1**, *k* represents the time index and 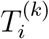 represents the total corona-positive cases corresponding the incubation period *τ*_i_, presented in **Fig. S1**.

The time-dependent model is the following set of coupled delay differential equations:

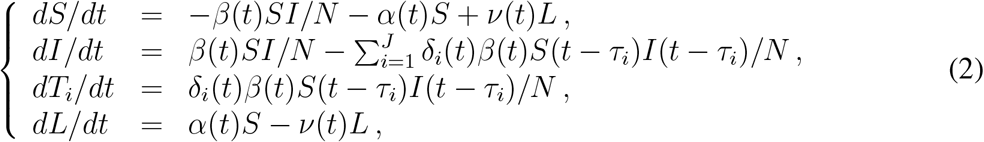

where *α*(*t*), *β*(*t*), *δ*_i_(*t*), for *i* = 1, ⋯, *J* and *ν*(*t*) are real positive parameters respectively modeling the rate of lockdown, the rate of infection, the rate of tested corona-positive corresponding the incubation period *τ*_i_ and the rate of ignoring lockdown, respectively. It follows from **Eqn. 2**, that for any *t*

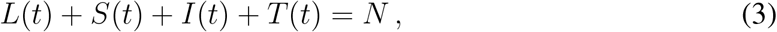

where *N* (constant) is the total population size.

We solve **Eqn. 2** using `matlab` inner-embedded program **dde23** with particular sets of model parameters. To solve the initial value problem **Eqn. 2**, in the interval [*t*_0_, *t*_1_], we consider *L*(*t*_0_), *S*(*t*_0_), *I*(*t*_0_) and *T* (*t*_0_) as follows:

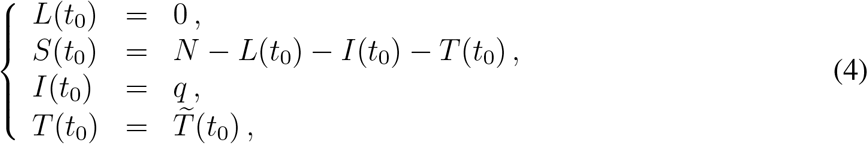

where 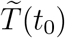 is the available data at time *t*_0_, and *q* is the initial value adjusting parameters. Initially, there is no lockdown individual so that we can consider *L*(*t*_0_) = 0.

### Parameter estimation of the model

We focus on the exponential growth phase of the COVID-19 epidemic in Canada; one can use the approach to estimate the incubation period distribution for any region affected by the infectious disease. The time resolved (daily updated) database (*1*) provides the number of total corona-positive cases. The optimal values of p(*t*) = (*q, α*(*t*), *β*(*t*), *δ*_1_(*t*), ⋯, *δ*_J_ (*t*), *ν*)^T^, that is the set of initial values and model parameters, is obtained by minimizing the root mean square error function *E*(p(*t*)), defined as

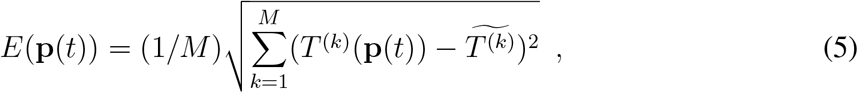

where 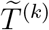 is the available data of total corona-positive cases on the particular *k*th day, and *T* ^(k)^ is the calculated results obtained from System **Eqn. 2**. The integer *M*, used in **Eqn. 5**, is the size of the data set. Due to the complexity of the error function, the minimization using the `matlab` function **fminsearch** requires a very large number of iterations.

### Numerical Experiment

In this section, we propose a detailed description of the computational procedure for the proposed model. On 23 January 2020, a 56-year old man admitted to Toronto hospital emergency department in Toronto with a new onset of fever and nonproductive cough, and returning from Wuhan, China, the day prior (*16, 24*). It is believed this is the first confirmed case of 2019-nCoV in Canada, and according to the government report, the novel coronavirus arrived on the Canadian coast on January 25, 2020, first reported case. The above information suggests that the start date of the current pandemic in Canada is possibly xsto be January 22, 2020. Additionally, some research studies reported that the estimation of the incubation period of COVID-19 is from 2 to 14 days (*2, 20*). As a consequence, in the present study we consider 14 delays, *τ*_1_ = 1 day, *τ*_2_ = 2 days, …, *τ*_14_ = 14 days. Here we consider a calculation of 276 days, from January 22, 2020 to October 23, 2020. We decompose the time domain of 276 days into two parts : the time domain splitter is in the interval where the first wave is slowed down and the “second wave” begins, i.e. the splitter is in the interphase of two different scenarios. In this goal, we can choose the parameters p(*t*) as

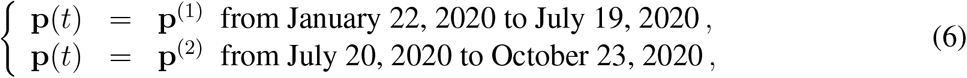

where 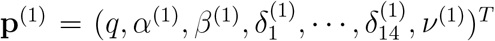 and 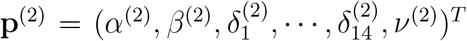 are some constants. The capability of an optimization package depends on the initial values of the parameters: for *q, α, β, ν* we consider any positive random number less than unity, where as a choice of **x** = (*δ*_1_, ⋯, *δ*_14_)^T^ is tricky. For this purpose, we consider a vector of 14 positive random numbers **x** such that *δ*_1_ *<* ⋯*< δ*_4_ *> δ*_5_ *> δ*_6_ *>* ⋯*> δ*_14_ and 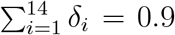. We observe, from numerous numerical experiments, the renormalization factor 0.9 works perfectly for the computation.

For a complete calculation, we run the `matlab` code twice. Firstly, we run the code for the period January 22, 2020 to July 19, 2020 to obtain the estimated value 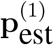 of p^(1)^, presented in **Table S1**, and the value of the error function 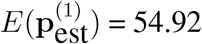. Then, we run the code for the entire period from January 22, 2020 to October 23, 2020, but using the estimated value 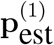 of p^(1)^for the interval January 22, 2020 to July 19, 2020, and obtain the estimated value 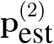 of p^(2)^ for the rest of the period, presented in **Table S1**, and the value of the error function 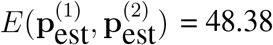, defined as

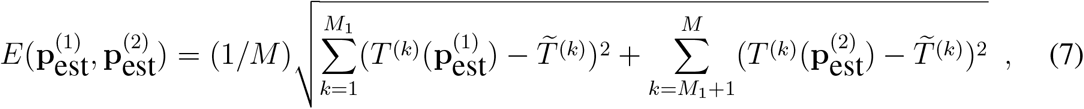

where *M*_1_ corresponds the date July 19, 2020.

## RESULTS

After estimating the model parameters with sufficiently small values of the error functions, we compare in **Fig. 1** total corona cases calculated with our model and the available data (*1*). This shows excellent agreement between the model results and the data. In **Fig. 2**, the confirmed cases 211,735 of 276 days are divided into fourteen groups. The *i*th compartment *T*_i_, defined in **Eqn. 1**, is the confirmed cases of 276 days corresponding to the incubation period of *i* day(s) for *i* = 1, 2, …, 14. In addition *T*_i_ is the frequency of the incubation period of *i* day(s), and using the bar chat, we obtain a mean incubation period of 6.89 days, a median of the incubation period of 6 days, 90th percentile of 11 days, 95th percentile of 12 days and 99th percentile of 13.5 days. The bar chat shows that mode of the incubation period is of 6 days, and there is a second peak for the incubation period at 10 days. However, the second peak is strongly dominated by the first. From the bar chat presented in **Fig. 2**, we can also obtain the probability densities of incubation period of the first *k* days during the epidemic, thanks to the total confirmed cases of the first *k* days starting on January 22, 2020. The probability densities 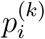 of the first *k* days and corresponding incubation period *i* days for *i* = 1, 2, …, 14 can be defined as

**Figure 1:**
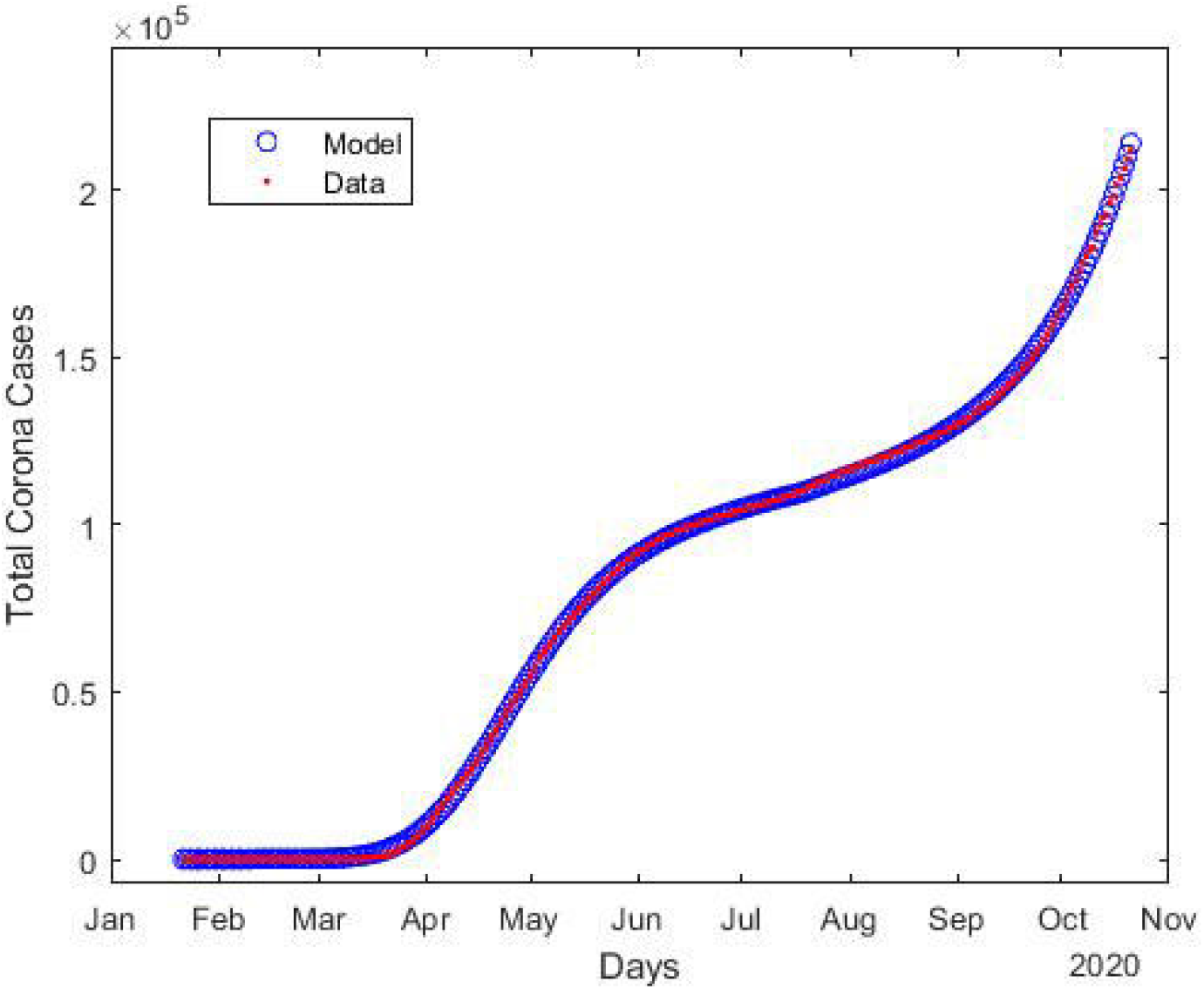
Estimation of the total number of confirmed coronavirus cases (*T*) compared to the available data (*1*). Here blue circles indicates the results obtained from model, and red line is the publicly available data (*1*).

**Figure 2:**
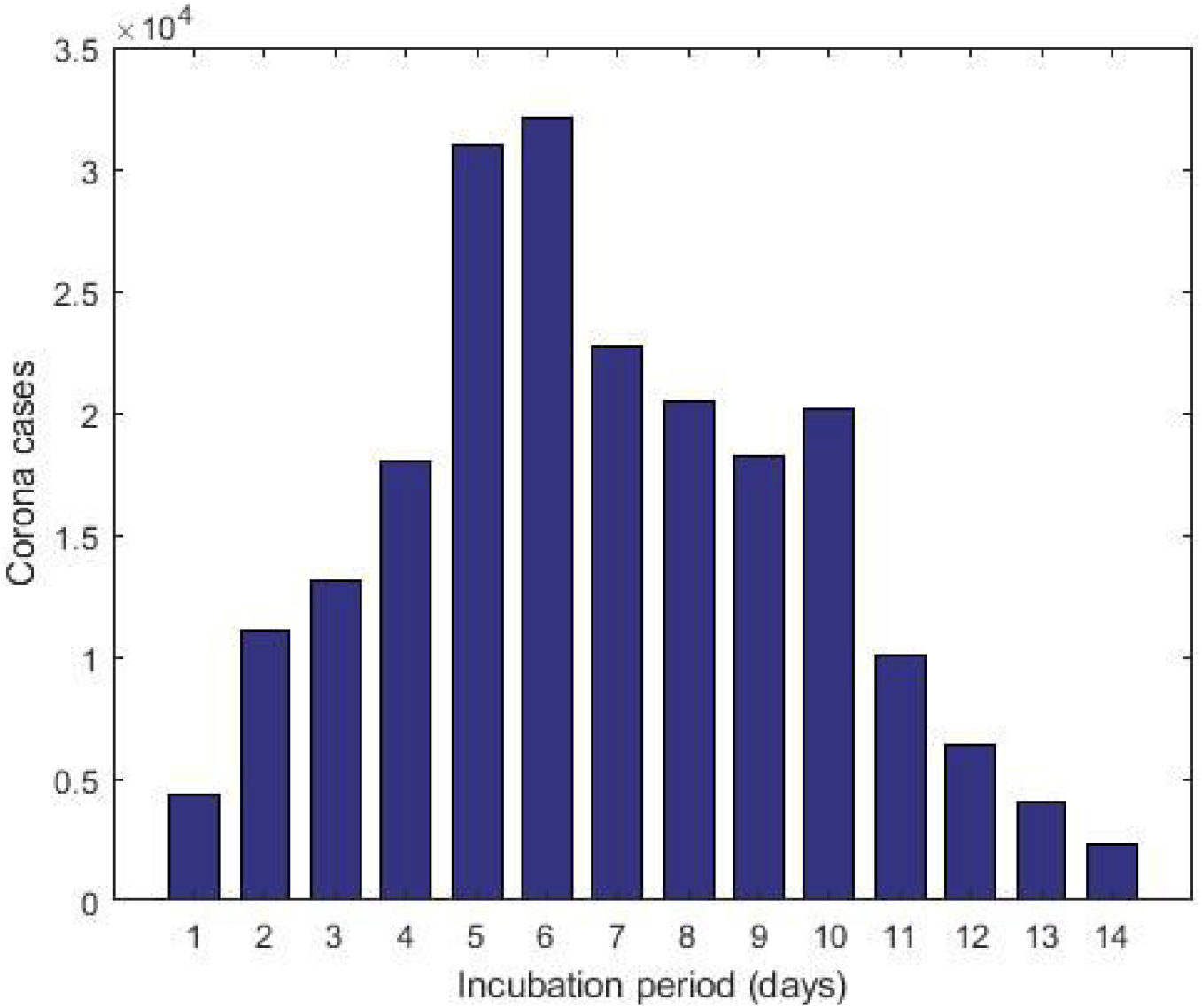
The cumulative data of confirmed corona cases as of October 23, 2020 is splitted into several incubation periods.

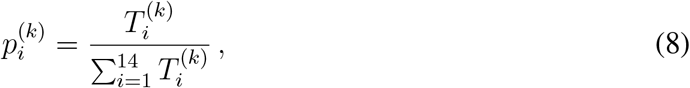

where 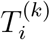 are defined in **Eqn. 1**. The probability densities 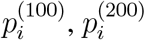 and 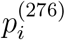 for *i* = 1, 2,, …, 14 are presented in **Fig. 3**. The density curves for the first 100, 200 and 276 days are similar, and the densities obey a remarkable configuration: for 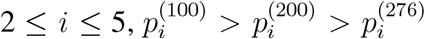 ^)^, for 7 ≤ *i* ≤ 9 and 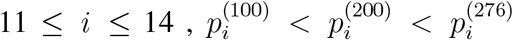 although the second peak, for *I* = 10, does not follow the decreasing-increasing convention. For the incubation period of 10 *days* 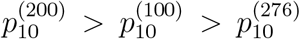 an oscillatory behaviour is observed. The descending pattern for 2 ≤ *i* ≤ 5 and the ascending order for 7 ≤ *i* ≤ 9 and 11 ≤ *i* ≤ 14 indicate that the mean incubation period is rising. Now, we fit the frequency data for the first 276 days, presented in the bar chat **Fig. 2**, with the lognormal distribution function and obtain the lognormal distribution parameters *µ* = 1.79 and *σ* = 0.52. **Fig. 4** shows the lognormal distribution function of the incubation period of the first 276 days and population size 211,735 with 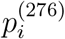 for *i* = 1, 2, …, 14. The estimated incubation period, obtained using lognormal distribution, has a mean of 6.74 (95% CI: 6.35 to 7.13), and the 90th percentile is 11.64 days (95% CI : 11.22 to 12.17). In addition, we focus on the distribution of the incubation period for a single day, October 23, 2020 which is the 276th day of the epidemic, with 2258 confirmed cases. The probability density 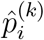 for a single day can be calculated as

**Figure 3:**
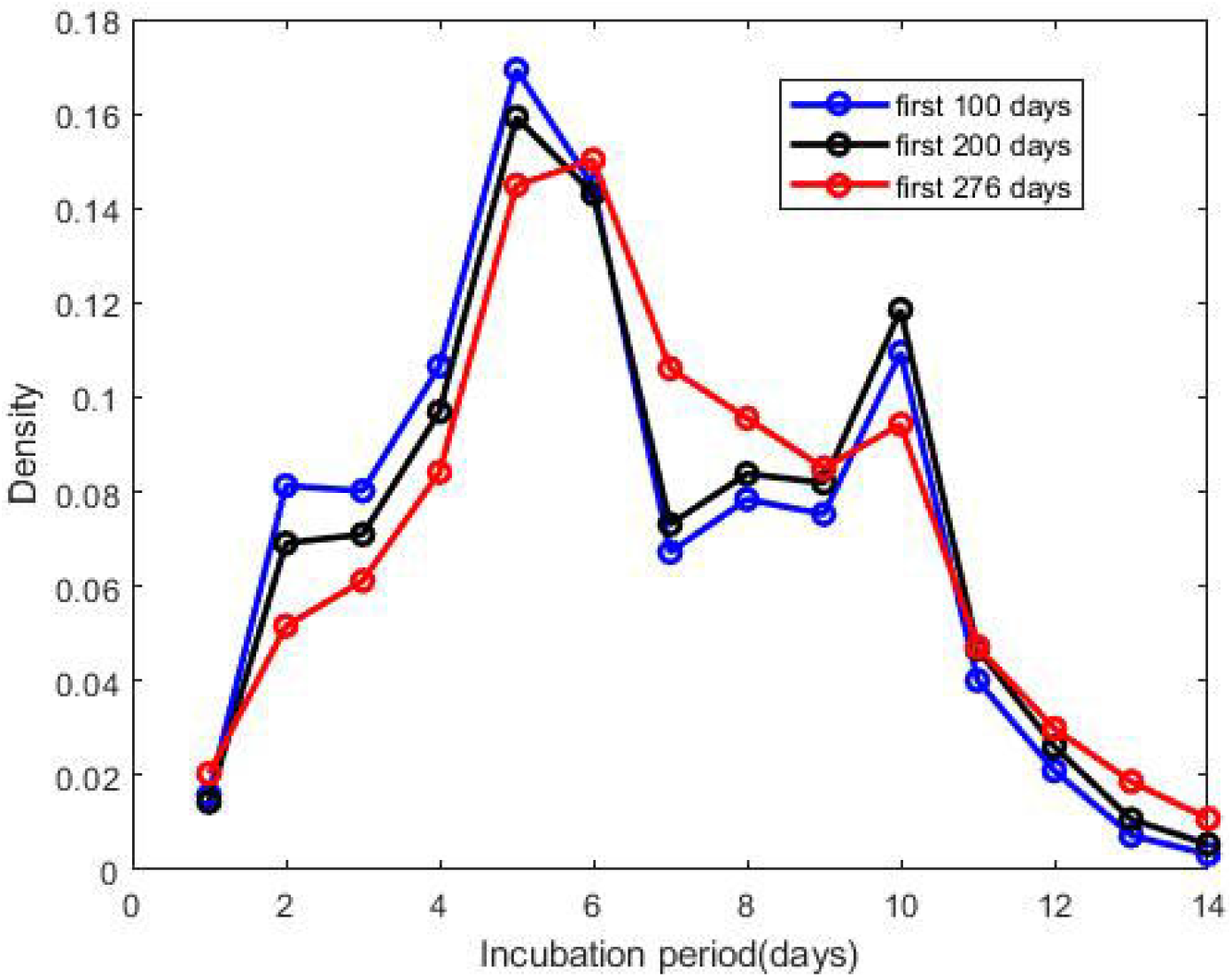
Probability densities of incubation period, presented in **Eqn. 8**. The ‘first 100 days’ indicates that the density of incubation period based on the cumulative data of the first 100 days during the epidemic starting from January 22, 2020 and similar for other two.

**Figure 4:**
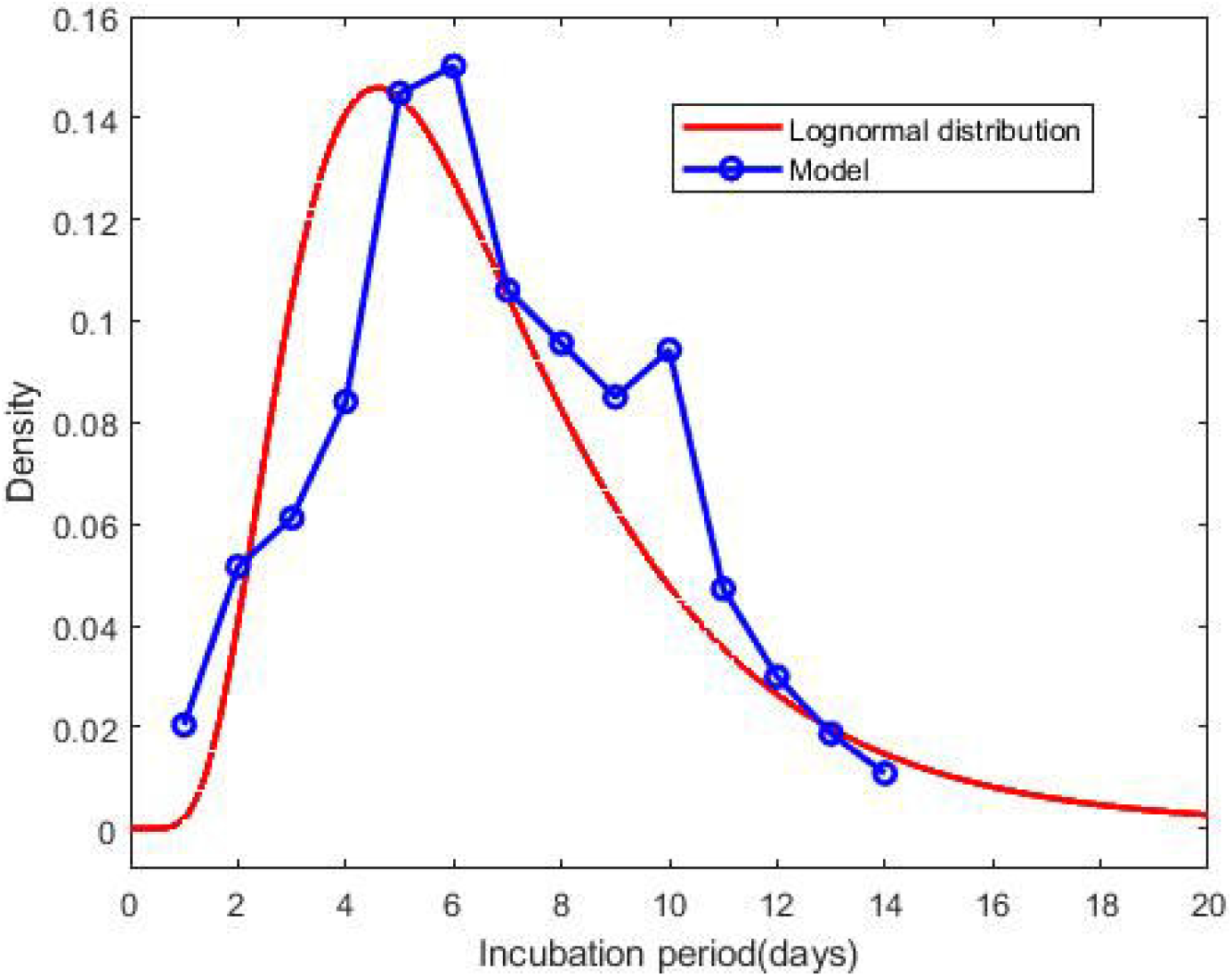
Probability density function of the lognormal distribution of the incubation period with *µ* = 1.79 and *σ* = 0.52. The result based on the total confirmed corona cases of 276 days. The blue circles indicate the densities obtained from the model calculation.

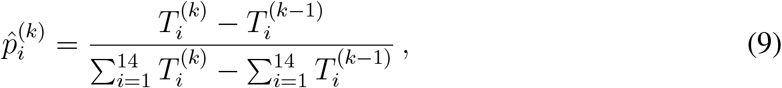

where 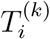 is defined in **Eqn. 1**. The estimated incubation period, obtained from frequency table of 276th day and population size of 2258, has a mean of 7.14 days, a median of 7 days, the 90th percentile of 11 days, 95th percentile of 12.5 days and 99th percentile of 14 days. We generate the lognormal distribution function from the 276th day’s frequency data and obtain the lognormal distribution parameters *µ* = 1.83 and *σ* = 0.53. **Fig. 5** shows the lognormal distribution of 276th day along with 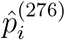. The estimated incubation period of 276th day, population size of 2258 and obtained using a lognormal distribution, has a mean of 6.98 days (95% CI: 6.41 to 7.55) and the 90th percentile of 12.29 days. A list of several studies along with the present calculation “Math.-Model” are presented in **Table 1**; we present the mean incubation period for the raw data as well as the lognormal distribution function. The list shows that our calculated data are closed the value reported by Ma *et. al*. (*15*) using a larger sample size of 587.

**Table 1:** A list of several studies along with sample size, mean and lognormal parameters *µ, σ*.

| Author | Data size | Mean (days) | $\mu$ | $\sigma$ |
| --- | --- | --- | --- | --- |
| Backer *et al.* (3) | 88 | 6.4 | 1.796 | 0.349 |
| Lauer *et al.* (11) | 181 | 5.5 | 1.621 | 0.418 |
| Li *et al.* (13) | 10 | 5.2 | 1.425 | 0.669 |
| Bi *et al.* (4) | 183 | 4.8 | 1.570 | 0.650 |
| Jiang *et al.* (9) | 40 | 4.9 | 1.530 | 0.464 |
| Linton *et al.* (14) | 158 | 5.6 | 1.611 | 0.472 |
| Zhang *et al.* (28) | 49 | 5.2 | 1.540 | 0.470 |
| Ma *et al.* (15) | 587 | 7.4 | 1.857 | 0.547 |
| Leung *et al.* (12) | 61 | 7.2 | 1.780 | 0.680 |
| McAloon *et al.* (17) | Meta | 5.8 | 1.63 | 0.50 |
| Jing *et al.* (23) | 1084 | 8.29 |  |  |
| Math Model (raw data) | 211,735 | 6.89 |  |  |
| Math Model (lognormal) | 211,735 | 6.7 | 1.788 | 0.520 |
| Math Model (raw data) | 2,587 | 7.17 |  |  |
| Math Model (lognormal) | 2,587 | 7.0 | 1.827 | 0.528 |

**Figure 5:**
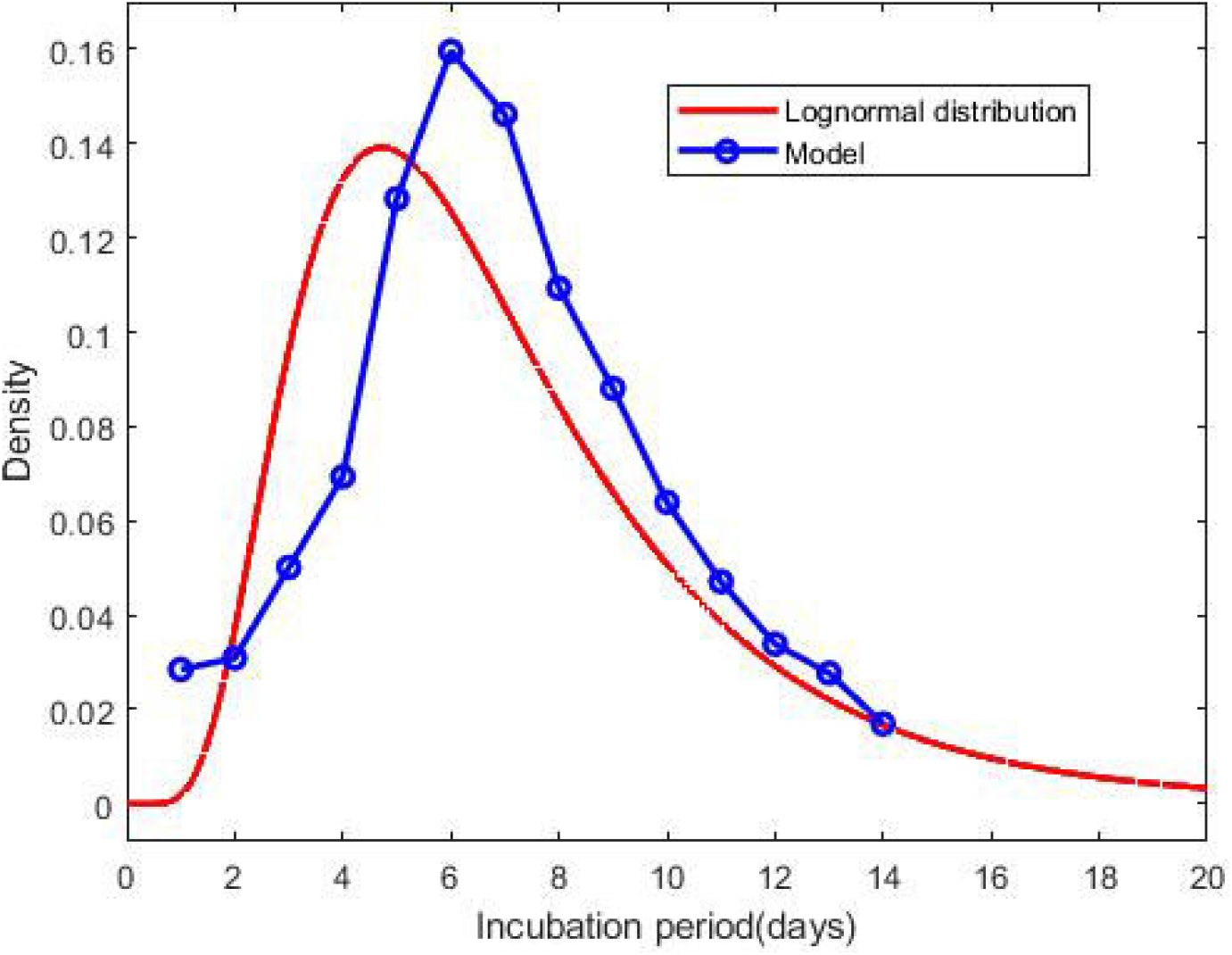
Probability density function of the lognormal distribution of the incubation period with *µ* = 1.83 and *σ* = 0.53. The result based on the confirmed corona cases of a particular day, October 23, 2020. The blue circles indicate the densities obtained from the model calculation.

## DISCUSSION

The calculated mean incubation period using two different ways, the raw data as well as the lognormal distribution are indeed closed, indicating that the raw data calculated using our mathematical model, are statistically significant for a lognormal distribution (statistical *p* value less than 0.001). It follows from the “Math.-Model” calculation, presented in **Table 1**, that the mean incubation period of 276th day, population size 2258, is greater than the mean incubation period of 276 days, population size 221,735 which demonstrates that the mean incubation period of COVID-19 is slightly increasing with time.

In this paper, we have derived a mathematical model based on a set of coupled delay differential equations, which was used to estimate the incubation period with good agreement with statistical works. Using the proposed model and publicly available data of confirmed cases, one could accurately estimate the incubation period in any region. We obtain the distribution of the incubation period from the population, so that it is better than any sample-dependent result. We have considered fourteen delays, but it is possible to consider an arbitrary number of delays. After estimating the model parameters, one can estimate the incubation period of confirmed cases over a long period, over a small time interval, and even over a single day. The present approach can be used with a large-scale computation to estimate the recovery period of COVID-19.

## Data Availability

Data obtained from publicly available Canadian web page.

https://resources-covid19canada.hub.arcgis.com/

## Funding

This research is supported by the Natural Sciences and Engineering Research Council of Canada (NSERC) and Mathematics of Information Technology and Complex Systems (MITACS) Ref. IT#19228.

## Author contributions

S.P. has derived the model, has developed the matlab code, has analyzed the calculated results, and has prepared all figures and tables. S.P. and E.L. have drafted the original article. Both authors have contributed to the editing of the article. Both authors have read and approved the final article.

## Competing interests

The authors declare that they have no competing interests.

## Data and materials availability

All data needed to evaluate the conclusions in the paper are present in the paper and/or the Supplementary Materials. Additional information related to this paper may be requested from the authors.

## Supplementary Materials

Figure 1 shows the compartmental based epidemic model; the set of coupled delay differential equations is generated from the model diagram. The estimated values of the model parameters are listed in Table 1. In Figure 2, the confirmed cases of first 100 days are divided into fourteen groups. The bar chat represents the frequency diagram of the incubation periods. From the bar chat, we also obtain the probability densities for first 100 days. The confirmed cases of first 200 days are bifurcated into fourteen groups, presented in Figure 3. In Figure 4 we report that the distribution of confirmed cases of October 23, 2020, a single day, for different incubation periods.

**Figure 1:**
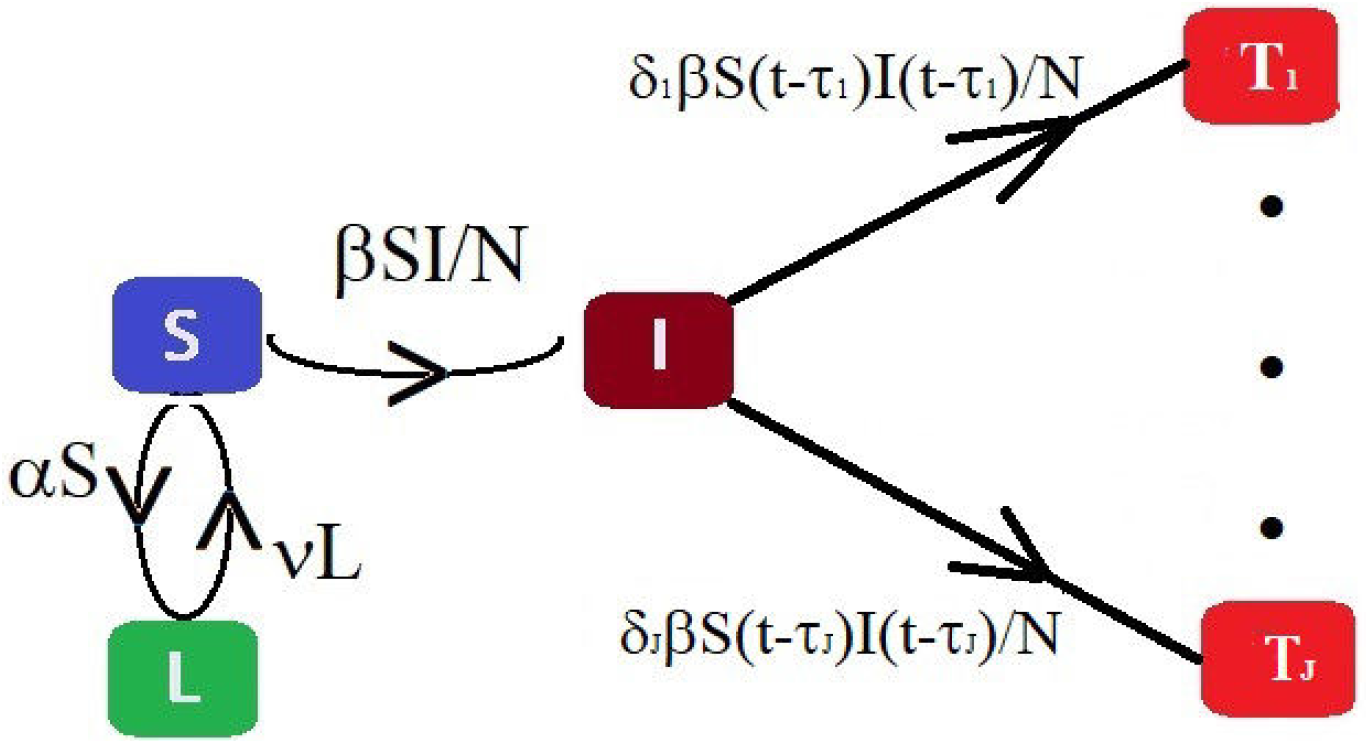
Schematic diagram of the compartmental based epidemic model, presented in Equation 2.

**Table 1:** The estimated values of the model parameters for two different time domains defined in Equation 6.

| Parameters | Estimated value | Parameters | Estimated value |
| --- | --- | --- | --- |
| $\alpha^{(1)}$ | 0.01376 | $\alpha^{(2)}$ | 0.00102 |
| $\beta^{(1)}$ | 0.44952 | $\beta^{(2)}$ | 0.73348 |
| $\delta_1^{(1)}$ | 0.01331 | $\delta_1^{(2)}$ | 0.02180 |
| $\delta_2^{(1)}$ | 0.07014 | $\delta_2^{(2)}$ | 0.02454 |
| $\delta_3^{(1)}$ | 0.07104 | $\delta_3^{(2)}$ | 0.04129 |
| $\delta_4^{(1)}$ | 0.09705 | $\delta_4^{(2)}$ | 0.05931 |
| $\delta_5^{(1)}$ | 0.15866 | $\delta_5^{(2)}$ | 0.11374 |
| $\delta_6^{(1)}$ | 0.13964 | $\delta_6^{(2)}$ | 0.14658 |
| $\delta_7^{(1)}$ | 0.06694 | $\delta_7^{(2)}$ | 0.13907 |
| $\delta_8^{(1)}$ | 0.08066 | $\delta_8^{(2)}$ | 0.10781 |
| $\delta_9^{(1)}$ | 0.08007 | $\delta_9^{(2)}$ | 0.08989 |
| $\delta_{10}^{(1)}$ | 0.12072 | $\delta_{10}^{(2)}$ | 0.06755 |
| $\delta_{11}^{(1)}$ | 0.04584 | $\delta_{11}^{(2)}$ | 0.05151 |
| $\delta_{12}^{(1)}$ | 0.02497 | $\delta_{12}^{(2)}$ | 0.03834 |
| $\delta_{13}^{(1)}$ | 0.00901 | $\delta_{13}^{(2)}$ | 0.03833 |
| $\delta_{14}^{(1)}$ | 0.00427 | $\delta_{14}^{(2)}$ | 0.02056 |
| $\nu^{(1)}$ | 0.00114 | $\nu^{(2)}$ | 0.00085 |

**Figure 2:**
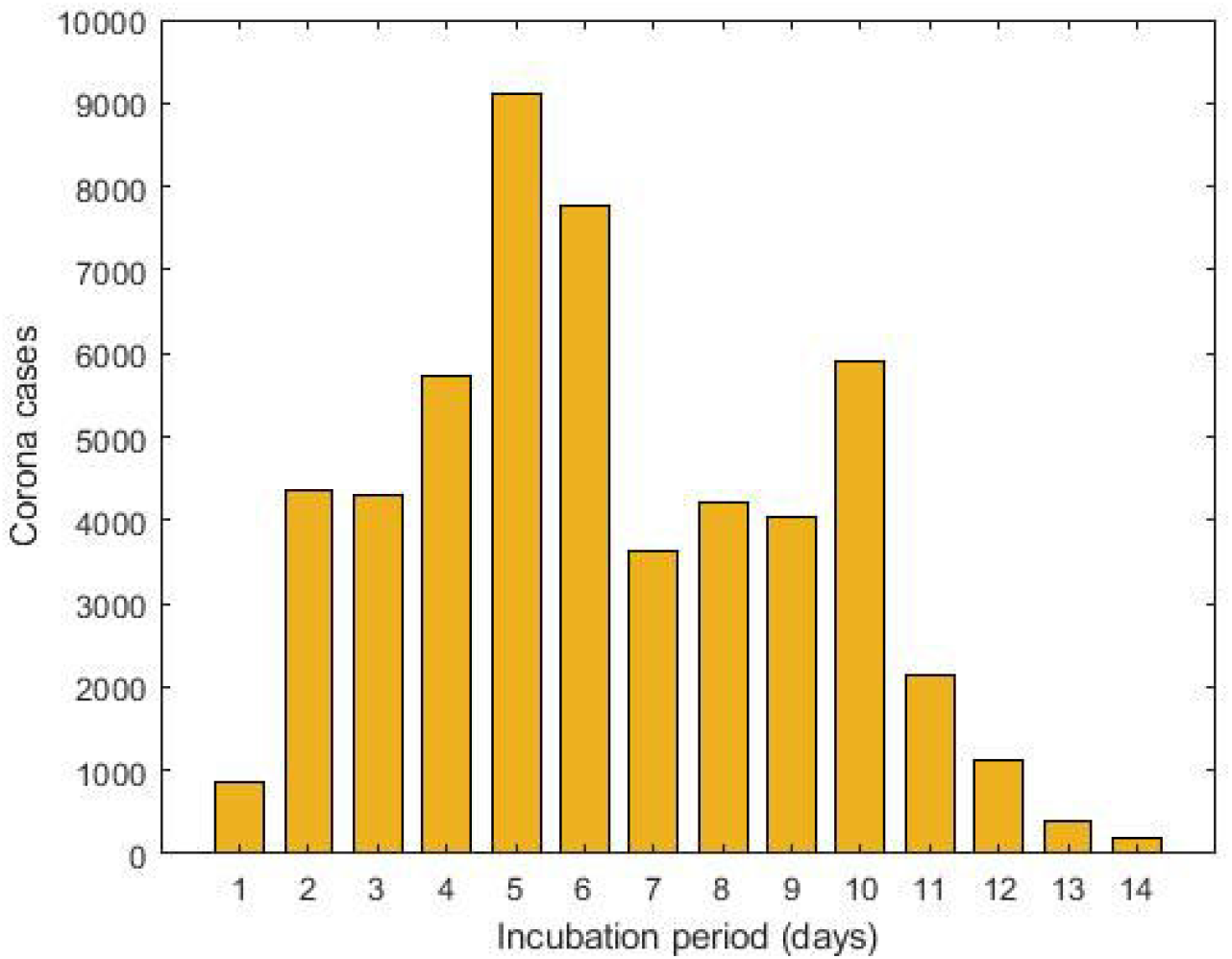
The cumulative data of confirmed corona cases for first 100 days is splitted into several incubation periods.

**Figure 3:**
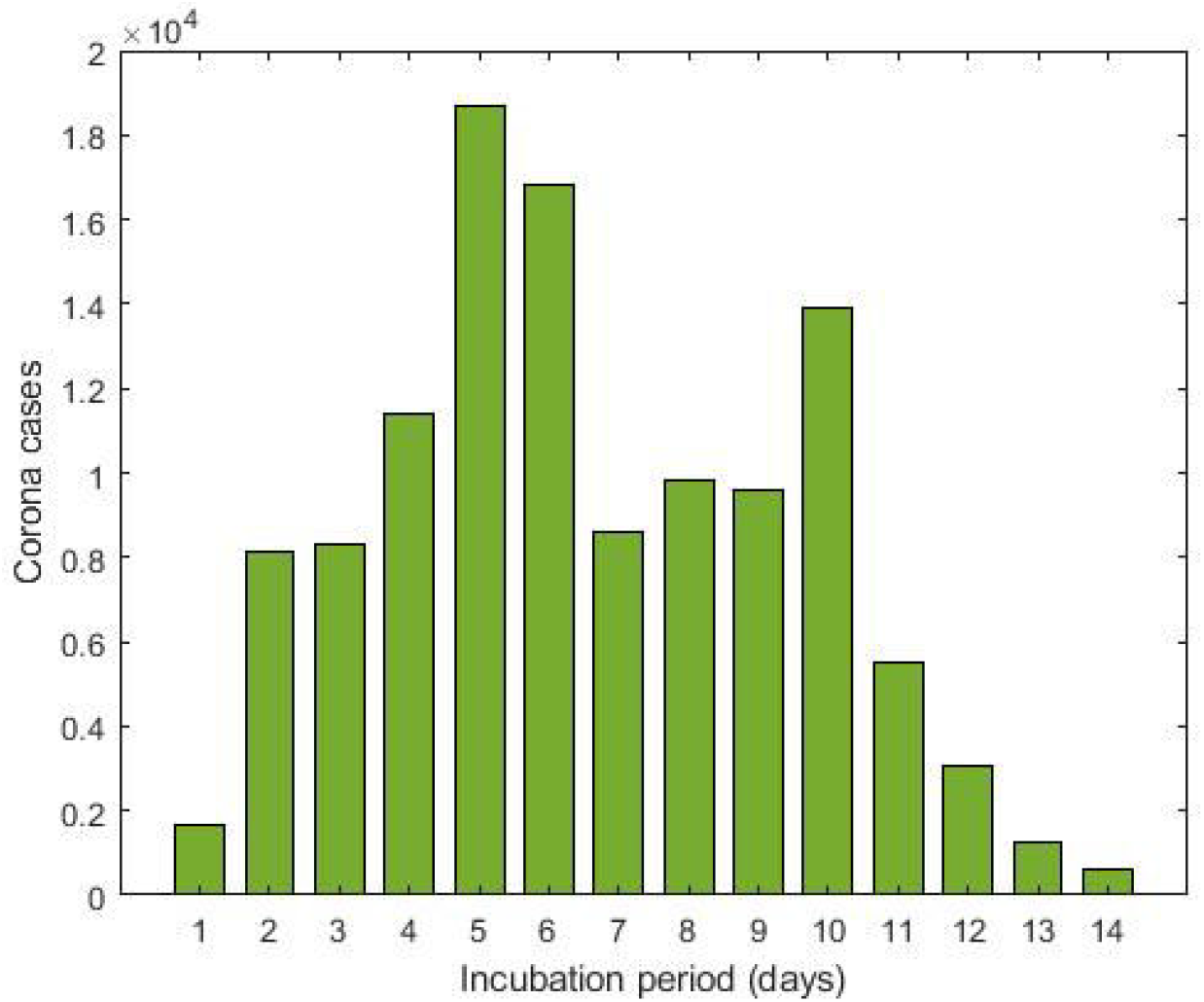
The cumulative data of confirmed corona cases for first 200 days is splitted into several incubation periods.

**Figure 4:**
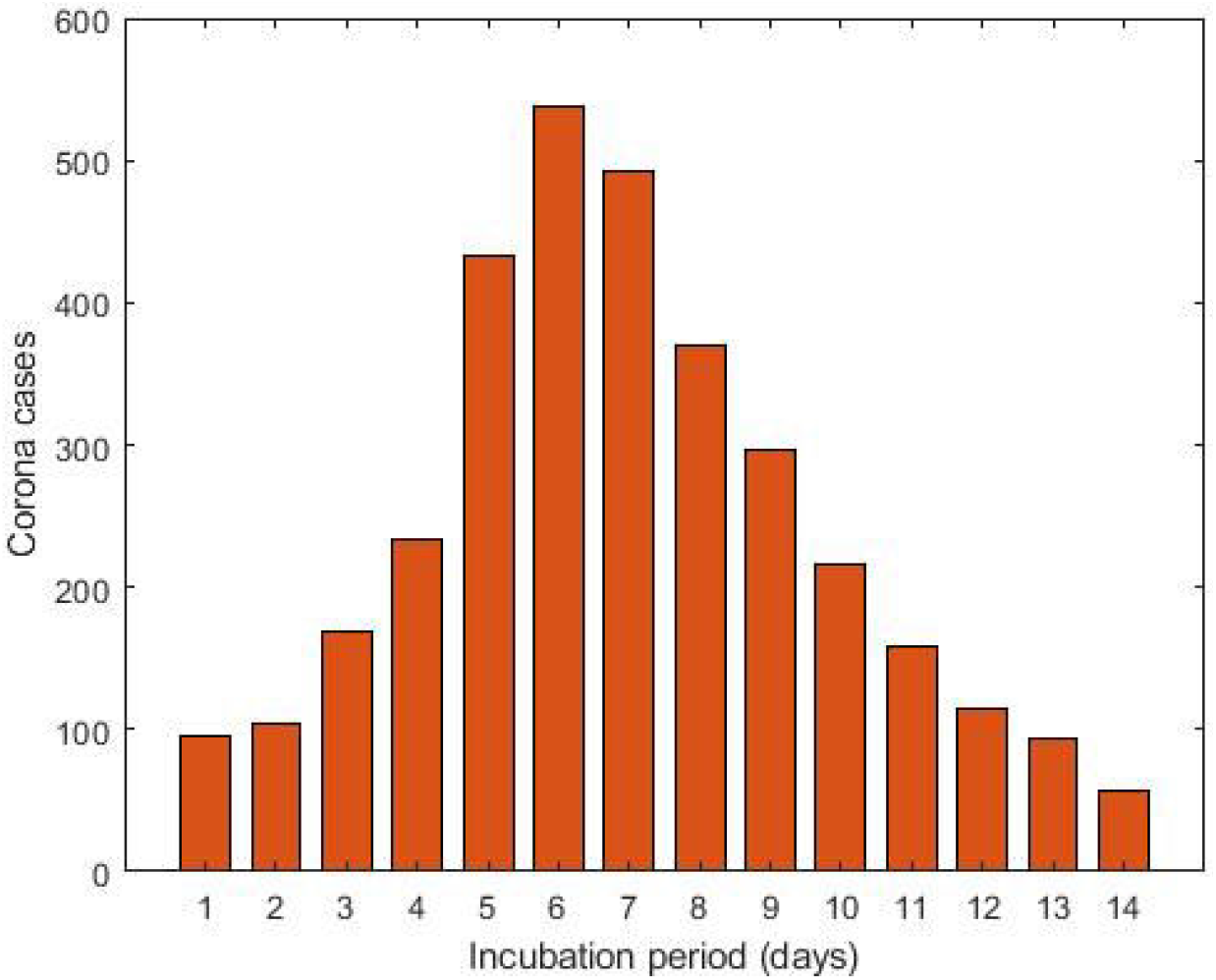
The confirmed corona cases as of October 23, 2020 is splitted into several incubation periods.

## Notes

### Competing Interest Statement

The authors have declared no competing interest.

## References and Notes

1. https://www.canada.ca/en/public-health/services/diseases/2019-novel-coronavirus-infection.html?topic=tilelink.x

2. https://www.worldometers.info/coronavirus/coronavirus-incubation-period/.

3. J. A. Backer, D. Klinkenberg, and J. Wallinga. Incubation period of 2019 novel coronavirus (2019-ncov) infections among travellers from Wuhan, China, 20–28 january 2020. Eurosurveillance, 25(5):2000062, 2020.

4. Q. Bi, Y. Wu, S. Mei, C. Ye, X. Zou, Z. Zhang, X. Liu, L. Wei, S. A. Truelove, T. Zhang, et al. Epidemiology and transmission of covid-19 in shenzhen China: Analysis of 391 cases and 1,286 of their close contacts. MedRxiv, 2020.

5. A. Bouchnita and A. Jebrane. A hybrid multi-scale model of covid-19 transmission dynamics to assess the potential of non-pharmaceutical interventions. Chaos, Solitons & Fractals, page 109941, 2020.

6. Y. Chen, J. Cheng, Y. Jiang, and K. Liu. A time delay dynamic system with external source for the local outbreak of 2019-ncov. Applicable Analysis, pages 1–12, 2020.

7. L. Dell’Anna. Solvable delay model for epidemic spreading: the case of covid-19 in italy. arXiv preprint arXiv:2003.13571, 2020.

8. A. Eshragh, S. Alizamir, P. Howley, and E. Stojanovski. Modeling the dynamics of the covid-19 population in australia: A probabilistic analysis. arXiv preprint arXiv:2005.12455, 2020.

9. X. Jiang, Y. Niu, X. Li, L. Li, W. Cai, Y. Chen, B. Liao, and E. Wang. Is a 14-day quarantine period optimal for effectively controlling coronavirus disease 2019 (covid-19)? medRxiv, 2020.

10. A. J. Kucharski, T. W. Russell, C. Diamond, Y. Liu, J. Edmunds, S. Funk, R. M. Eggo, F. Sun, M. Jit, J. D. Munday, et al. Early dynamics of transmission and control of covid-19: a mathematical modelling study. The lancet infectious diseases, 2020.

11. S. A. Lauer, K. H. Grantz, Q. Bi, F. K. Jones, Q. Zheng, H. R. Meredith, A. S. Azman, N. G. Reich, and J. Lessler. The incubation period of coronavirus disease 2019 (covid-19) from publicly reported confirmed cases: estimation and application. Annals of internal medicine, 172(9):577–582, 2020.

12. C. Leung. The difference in the incubation period of 2019 novel coronavirus (sars-cov-2) infection between travelers to Hubei and nontravelers: The need for a longer quarantine period. Infection Control & Hospital Epidemiology, 41(5):594–596, 2020.

13. Q. Li, X. Guan, P. Wu, X. Wang, L. Zhou, Y. Tong, R. Ren, K. S. M. Leung, E. H. Y. Lau, J. Y. Wong, et al. Early transmission dynamics in Wuhan, China, of novel coronavirus– infected pneumonia. New England Journal of Medicine, 2020.

14. N. M. Linton, T. Kobayashi, Y. Yang, K. Hayashi, A. R. Akhmetzhanov, S.-M. Jung, B. Yuan, R. Kinoshita, and H. Nishiura. Incubation period and other epidemiological characteristics of 2019 novel coronavirus infections with right truncation: a statistical analysis of publicly available case data. Journal of clinical medicine, 9(2):538, 2020.

15. S. Ma, J. Zhang, M. Zeng, Q. Yun, W. Guo, Y. Zheng, S. Zhao, M. H. Wang, and Z. Yang. Epidemiological parameters of coronavirus disease 2019: a pooled analysis of publicly reported individual data of 1155 cases from seven countries. Medrxiv, 2020.

16. X. Marchand-Senécal, R. Kozak, S. Mubareka, N. Salt, J. B. Gubbay, A. Eshaghi, V. Allen, Y. Li, N. Bastien, M. Gilmour, et al. Diagnosis and management of first case of covid-19 in Canada: Lessons applied from sars. Clinical Infectious Diseases, 2020.

17. C. McAloon, Á . Collins, K. Hunt, A. Barber, A. W. Byrne, F. Butler, M. Casey, J. Griffin, E. Lane, D. McEvoy, et al. Incubation period of covid-19: a rapid systematic review and meta-analysis of observational research. BMJ open, 10(8):e039652, 2020.

18. J. Menendez. Elementary time-delay dynamics of covid-19 disease. medRxiv, 2020.

19. K. Y. Ng and M. M. Gui. Covid-19: Development of a robust mathematical model and simulation package with consideration for ageing population and time delay for control action and resusceptibility. Physica D: Nonlinear Phenomena, page 132599, 2020.

20. World Health Organization. Rolling updates on coronavirus disease (covid-19).[cited 2020 april 14] available at: https://www. who. int/emergencies/diseases/novel-coronavirus-2019/events-as-they-happen. 2020.

21. S. Paul and E. Lorin. Lockdown: a non-pharmaceutical policy to prevent the spread of covid-19. mathematical modeling and computation. 2020.

22. Y. Qaddura and N. Mavinga. Analysis of a vector-borne diseases model with a two-lag delay differential equation. The North Carolina Journal of Mathematics and Statistics, 4:12–28, 2018.

23. J. Qin, C. You, Q. Lin, T. Hu, S. Yu, and X.-H. Zhou. Estimation of incubation period distribution of covid-19 using disease onset forward time: a novel cross-sectional and forward follow-up study. medRxiv, 2020.

24. W. K. Silverstein, L. Stroud, G. E. Cleghorn, and J. A. Leis. First imported case of 2019 novel coronavirus in Canada, presenting as mild pneumonia. The Lancet, 395(10225):734, 2020.

25. W. Y. T. Tan, L. Y. Wong, Y. S. Leo, and M. P. H. S. Toh. Does incubation period of covid-19 vary with age? a study of epidemiologically linked cases in singapore. Epidemiology & Infection, 148, 2020.

26. C. P. Vyasarayani and A. Chatterjee. New approximations, and policy implications, from a delayed dynamic model of a fast pandemic. arXiv preprint arXiv:2004.03878, 2020.

27. Z. Yin, Y. Yu, and Z. Lu. Stability analysis of an age-structured seirs model with time delay. Mathematics, 8(3):455, 2020.

28. J. Zhang, M. Litvinova, W. Wang, Y. Wang, X. Deng, X. Chen, M. Li, W. Zheng, L. Yi, X. Chen, et al. Evolving epidemiology of novel coronavirus diseases 2019 and possible interruption of local transmission outside Hubei province in China: a descriptive and modeling study. MedRxiv, 2020.

